# Microbiome signature in wastewater is predictive of COVID-19 case rates

**DOI:** 10.64898/2026.09.16.26363250

**Authors:** Ty A. Schroeder, Christopher M. Basting, Erik Swanson, Melisa Bailey, Shannon R. Champeau, Adrian Velez, Jing Li, Courtney Broedlow, Timothy W. Schacker, Mark J. Osborn, Nichole R. Klatt

## Abstract

Surveillance of SARS-CoV-2 through wastewater proved crucial during the height of the pandemic for estimating community COVID-19 cases and continues to be useful for monitoring this and other pathogens. Gastrointestinal symptoms of acute COVID-19 have been well characterized, with demonstrated significant effects on the respiratory, oral, and gut microbiomes. With this understanding, we sought to characterize the wastewater microbiome in the context of COVID-19 rates, identify associations with the wastewater SARS-CoV-2 viral loads, and apply modeling to assess the ability of the wastewater microbiome to predict COVID- 19 case rate. We observed numerous microbes associated with case rate including genera that are known to be depleted in vivo during SARS-CoV-2 infection such as keystone SCFA-producers such as *Blautia, Dorea*, and *Akkermansia* with implicated pathobionts *Ruminococcus gnavus* group and *Ruminococcus torques* group that expand. Non-human origin bacteria including *Uruburuella* and *Stenoxybacter* were also strongly associated with community case rates. Our predictive models highlighted a combination between bacteria and SARS-CoV-2 gene copies as an effective combination of covariates to estimate present cases, however microbiome covariates alone proved modestly more useful in predicting future case rates. These results highlight the heavy influence of local ecosystem and urbanicity in defining microbial composition of WWTPs. Alterations in the fecal microbiome from acute COVID-19 infection are detectable in wastewater and strongly associated with case rate, even in low-population areas. Our predictive models underscore the utility of the wastewater microbiome in understanding public health burden of COVID-19 and possibly other infectious diseases.

**Importance:** Ongoing COVID-19 infections and persistent symptoms continue to contribute to an expanding public health burden that is challenging to track. While overall transmission and symptom severity have tapered in acute infection leading to reduced hospitalizations, thanks in part due to widespread vaccination efforts, predicting cases provides health care units time to mobilize and adapt. This study supports a growing body of evidence that the host gut microbiome is heavily altered from infection and highlights that this consistent alteration can be harnessed to detect microbial shifts in wastewater. Here, we used the wastewater bacterial microbiome to predict community cases weeks ahead of when cases were reported with accuracy matching current standard techniques. This work furthers our understanding of COVID-19’s impact on the gut microbiome, advances current wastewater-based epidemiology strategies, offers a new pool of data to better track community infections, and may support surveillance of other pathogens and novel, emerging outbreaks.

## Introduction

Wastewater-based epidemiology measures biological or chemical signals in water collected from sanitary sewers to offer insights about human populations, such as quantification of antibiotic resistance genes or disease prevalence in a community^1,2^. The surveillance of wastewater is also used to monitor levels of pathogens such as norovirus or poliovirus and was quickly adopted during the COVID-19 pandemic to estimate infection rates and identify emerging genetic variants^3,4^. While nasopharyngeal (NP) and oropharyngeal (OP) swabs are the most common method for identifying SARS-CoV-2 infection in individuals, wastewater surveillance enables infection estimates on a community scale, even when community members don’t have access to testing, choose not to seek testing or are asymptomatic. Even when NP and OP tests are available, their accuracy is not absolute as has been demonstrated by wastewater surveillance. In one study of repatriation flights carrying Australian citizens, wastewater collected from 24 of 37 flights tested positive for SARS-CoV-2 despite all passengers testing negative using deep nasal and oropharyngeal reverse-transcription polymerase chain reaction (RT-PCR) swab within 48 hours of flight. During the 14-day mandatory quarantine period following flight, 112 COVID-19 cases were detected demonstrating a discrepancy between swab and wastewater testing^5^.

This gap may be related to differences in viral replication between respiratory and gastrointestinal (GI) tissues, both of which highly express the angiotensin converting ezyme-2 (ACE2), the host receptor SARS-CoV-2 binds to and uses to enter cells. While primarily classified as a respiratory illness, gastrointestinal symptoms are common among patients with COVID-19 with some patients experiencing GI symptoms in the absence of respiratory symptoms^6^. Epidemiological evidence has yet to demonstrate widespread fecal-oral transmission of SARS-CoV-2^7^, suggesting an alternative route such as swallowed saliva or respiratory secretions. Coronaviruses are capable of surviving digestion, invading and replicating inside of intestinal M cells, and causing inflammation, gut barrier damage, diarrhea, and fecal viral shedding^8,9^. The length of GI shedding varies based on disease severity but may last long after respiratory shedding ends at about 17 days^10–12^. This GI viral shedding is a major component of the signal detected by wastewater surveillance^4^; however, viral particles may only be one type of microbial wastewater signal.

During active SARS-CoV-2 infection, the GI microbiome has demonstrated reduced alpha diversity and is enriched with opportunistic pathogens such as *Streptococcus anginosus*^13^ and *Pseudomonas spp*. ^14^. Not only are pathogens increased but the gut is depleted of essential short-chain fatty acid (SCFA)-producing beneficial microbes such as *Bifidobacterium, Blautia, and Faecalibacterium*^15^. Some of these perturbations persist after active infection has ended, especially in post-acute COVID syndrome patients (Long COVID), who continue to demonstrate depleted SCFA-producers such as *F. prausnitzii and Bacteroides thetaiotaomicron*^16^. These changes measured on an individual scale can be applied on a community-scale with wastewater surveillance, as was demonstrated in one study in which changes in the 16S microbiome profile of wastewater were associated with SARS-CoV-2 particles across dormitories on a campus^17^. The authors report a positive relationship between high SARS-CoV-2 concentrations and certain genera that are potential pathogens and an overall increase in diversity measures in SARS-CoV-2 positive samples. Potential pathogenic taxa such as *Arcobacter, Aeromonas,* and *Laribacter* were strongly associated with samples positive for SARS-CoV-2. Another study investigating 16S sequencing wastewater during the pandemic found a core set of phyla common to the 48 samples composed of Firmicutes, Proteobacteria, Bacteroidetes, and Actinobacteria. At the family level, *Lachnospiraceae*, *Bacteroidaceae*, and *Ruminoccoccacea* were the most abundant taxa. The authors discuss how wastewater surveillance can be used as an early detection system for pandemics and as a means to characterize common co-infections^18^.

In the present study, we report findings from an investigation of wastewater 16S sequencing data compared to matching SARS-CoV-2 viral loads and COVID-19 case rates from 13 wastewater treatment sites around the state of Minnesota between the months of January and November 2021. This investigation seeks to extend what is known about 16S wastewater surveillance during the COVID-19 pandemic with analysis of samples with a state-wide geographic and longitudinal distribution and use computational modeling to determine the wastewater microbiome’s efficacy in COVID-19 case rate determination.

## Methods

### Study Design

We collected samples from 13 wastewater treatment plants across the state of Minnesota (**Figure 1**) and the sewer shed population that each of these plants serve was estimated by the geographic proximity to different facilities (**Table 1**). Individual samples were a 24-hour composite of influent from each site that were collected weekly from January to November 2021.

**Figure 1.**
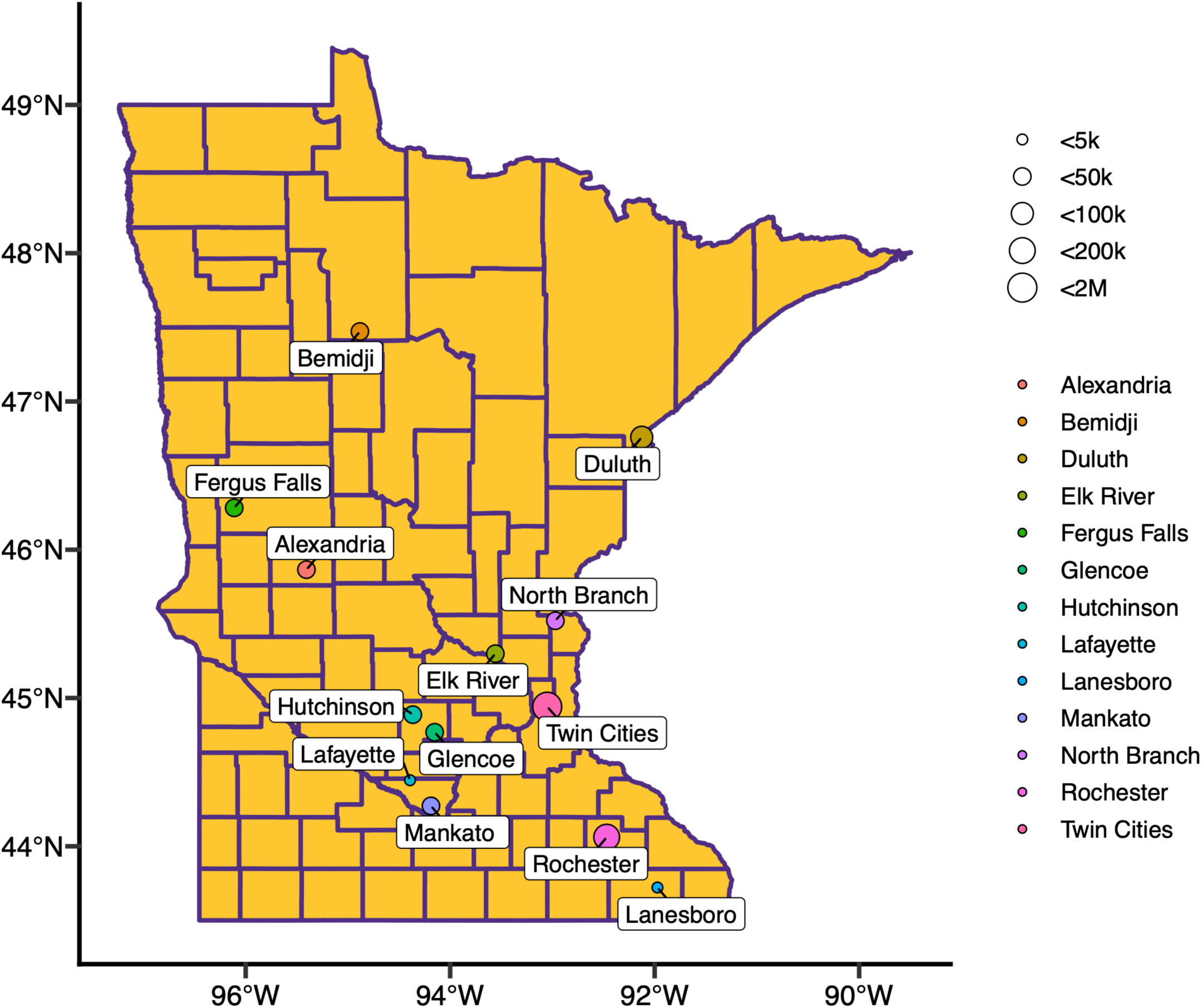
Location of WWTPs across Minnesota, USA. All wastewater facilities are located in Minnesota but varied by county. Number indicates the identity of each facility and dot size represents the respective sewershed population served.

**Table 1.**
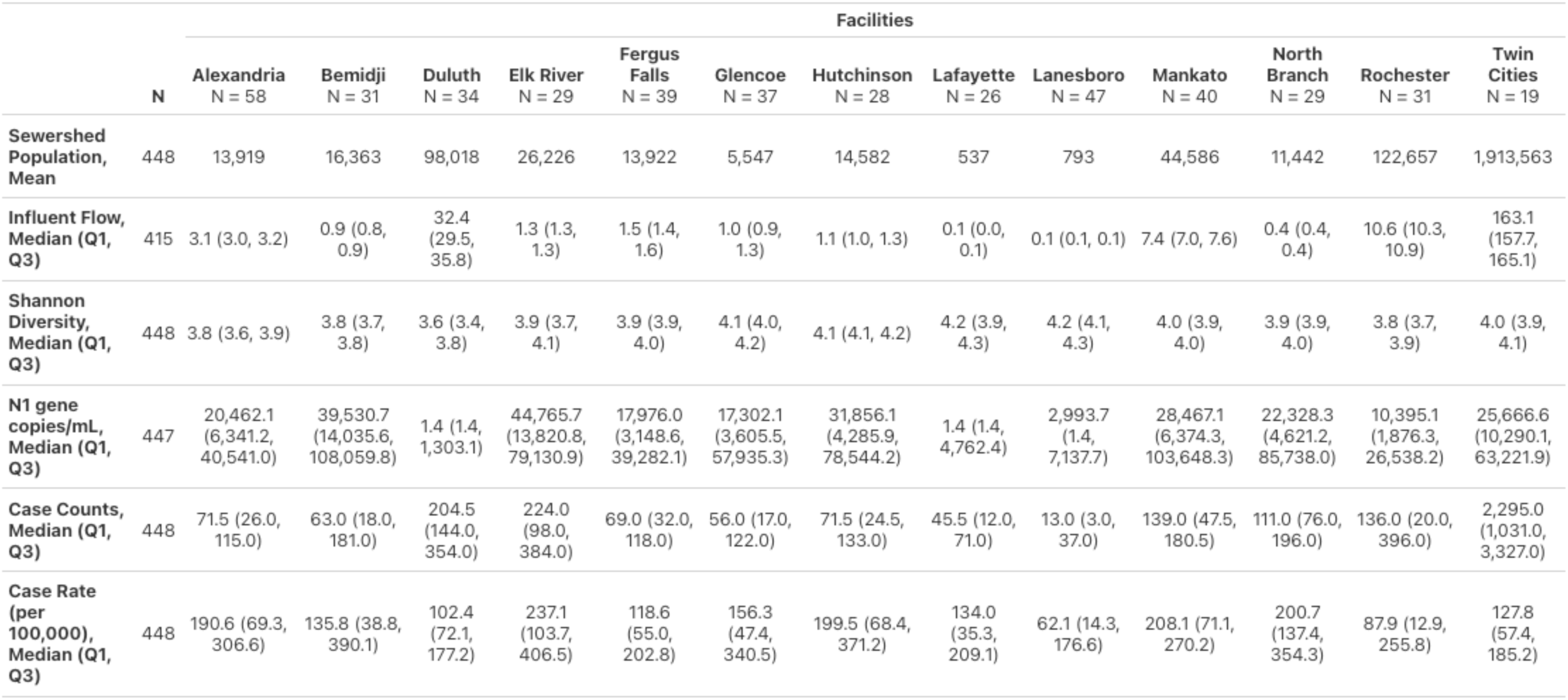
WWTP summary statistics across study collection period.

SARS-CoV-2 nucleocapsid (N)1 and N2 gene copies were measured from wastewater using qRT-PCR to calculate the viral copies per liter in wastewater as previously described^19^. Case counts and case rates were acquired from the Minnesota Department of Health (MDH) and detailed by county for the location of each respective wastewater treatment facility. Case counts and case rates were averaged by collection week to match the influent sample collections frequency. Daily high temperatures measured at stations closest to each facility were accessed from the National Oceanic and Atmospheric Administration (NOAA).

### 16S Sequencing

For each sample, 15 mL of wastewater was centrifuged at 4,500 x g for 30 minutes, supernatant decanted, and the pellet was resuspended in 1mL of DNA/RNA Shield (Zymo Research, Irving, CA). Microbial DNA was extracted using a modified version of the Qiagen DNeasy 96 PowerSoil Pro extraction kit (Qiagen, Hilden, Germany). Briefly, 250 µL of the resuspended sample was centrifuged at 16,000 × g for 1 minute. The supernatant was retained and the pellet was resuspended in CD1 buffer and carried through the standard lysis protocol. Prior to CD3 buffer addition, the retained supernatant was combined with the lysate and the volume of CD3 buffer was adjusted proportionally to accommodate the increased volume. The combined lysate was then processed according to the manufacturer’s instructions. Extracted DNA was submitted to the University of Minnesota Genomics Center for V4 16S rRNA sequencing on an Illumina MiSeq with an average sequencing depth of 37,523 reads per sample. *Cutadapt* (version 4.4)^20^ was used to remove variable region primers from the raw sequences. Sequences were then trimmed, denoised, and merged via the *dada2* package (version 1.32.0)^21^, keeping only those sequences that are of the expected length. Taxonomy was assigned to the amplicons sequence variants (ASVs) using *SILVA* (version 138)^22^. ASVs that related to chloroplast or mitochondria and uncharacterized at the phylum level were removed. Taxa were filtered to require a minimum of 25 counts in a given sample and present in at least 10% (45/448 observations) of samples to be included to remove noise from low abundant taxa using *microViz* (version 0.12.6)^23^, resulting in 310 taxa.

### Microbiome analysis

The methods to characterize the microbiome present in different facilities individually and in relation to one another using various metrics. For species evenness and richness within samples, alpha diversity indices (Observed, Shannon, and Simpson) were calculated for each sample using the *phyloseq* package (1.48.0)^24^. To test for significant differences in alpha diversity between the facilities, estimated marginal means were calculated using the *emmeans* package (1.10.2)^25^ by predicting Shannon diversity by facility and accounting for lack of independence between observations within facilities by including collection date by facility from the corCAR1 function within the gls function from the *nlme* package (3.1-169)^26^. Beta diversity was calculated from a dissimilarity matrix between samples based on Bray-Curtis distance also using *phyloseq* and then plotted in a principal coordinate analysis (PCoA) to visualize using the *microViz* package; centroids representing the average dissimilarity value of each facility were added to compare between facilities. Principal component analysis was calculated using *factoextra* (version 2.0.0) and PERMANOVA between facilities accounted for repeated measures by including case rate and an interaction of time and Facility using the adonis2 function in *vegan* (version 2.7-3)^27^. Bacteria relative abundance was transformed using robust center log ratio (RLCR) via the transform function within the *microbiome* (version 1.26.0)^28^ package prior to PCA. A Mantel test was conducted using *vegan* on the Euclidean distance between PCA centroids and Haversine distance of the physical location of facilities calculated from *geosphere* (version 1.6-8)^29^. Differential abundance of bacterial taxa with COVID-19 case rate as a fixed effect and facility as a random effect was conducted using *MaAsLin2* (1.18.0)^30^ to explore relationships between taxa and case rates.

### Statistical Analysis

All statistical analysis and data processing was conducted in R (4.4.0)^31^. Basic summary statistics were summarized and visualized using the *gtsummary* package (2.5.0)^32^. Repeated measure correlations on study covariates and microbiome measures were conducted using *rmcorr* (version 0.7.0)^33^. We assessed the predictive ability of various COVID-19 and microbiome measures on case rate by county using a leave-group-out cross validation (LGOCV) approach. Bacterial relative abundance data was center log ratio (CLR) transformed with a pseudocount of 1 prior to inclusion in models. Within each iteration, a single facility was left out and used as the test/unseen data and the remaining data points were a training set. Within these iterations, we assessed the predictive ability of various COVID-19 and microbiome measures on case rate by county with the final models using SARS-CoV-2 N1 gene copies/mL normalized by the daily influent rate means within facilities and relative abundance of selected bacteria. A generalized linear mixed model least-absolute-shrinkage and selection operator (glmmLASSO) was used to reduce the number of taxa that are applied to a model. An optimal penalty lambda value was chosen by step-wise testing a range of values and finding the lowest Bayesian information criterion (BIC) within the *glmmLasso* package (1.6.3)^34^. Individual mixed models were constructed in three kinds: normalized N1 gene copies/mL alone, feature selected bacteria alone, or a combination of the feature selected taxa and N1 gene copies. All models adjusted for the effect of seasonality by including temperature, a likely confounder, as a fixed effect and facility as a random effect. For each type of model, the county-level case rate was unchanged or had a 1- and 2-week lead effectively testing how effective covariates were at predicting the case rate 1 or 2 weeks in the future. Model metrics included the magnitude of error from observed values (root mean squared error; RMSE and mean absolute error; MAE) and then compared by analysis of variance (ANOVA) adjusting for inclusion of repeated data between model types, calculated with the base R *stats* package. Model metrics and all other visualizations were made using *ggplot2* (3.5.1)^35^.

## Results

### Characterization of wastewater treatment plants by microbiome measures

Although all wastewater facilities were located within Minnesota, they spanned a wide geographic range with substantial variation in rurality and demographics. Because there were disparities in sewer shed populations, we wanted to evaluate differences in the microbiome between facilities. The top ten abundant taxa were largely characterized by non-human origin bacteria such as *Leptotrichia* and *Acinetobacter* (**Supplementary Figure 1**). Abrupt changes in microbial composition appeared to be facility dependent with locations such as Duluth experiencing several sharp compositional perturbations characterized by changes in *Pseudoarcobacter, Psychrobacter*, and *Cloacibacterium*. Simple linear mixed models evaluating relationships of these bacteria with case count or temperature in Duluth indicated that *Pseudarcobacter* was significantly and negatively associated with case counts but not temperature (*p* = 0.00156 and *p* = 0.286, respectively) while *Psychrobacter* and *Cloacibacterium* were associated with temperature increases (*p* = 0.991, *p* = 0.0365; *p* = 0.548, *p* = 0.0166, respectively).

We next evaluated the richness and evenness of the facilities by calculating the alpha diversity and modeling the influence that each facility had on Shannon diversity. The resulting pairwise comparisons between facilities are shown in **Figure 2**. Most of the facilities contrasts were significantly different from one another, indicating that the richness and evenness of bacteria in wastewater varies greatly depending on location. Duluth had significant contrasts with all other facilities such as Lanesboro and Twin Cities metro (*q* < 0.001), owed to a smaller alpha diversity emmean value with negative *t*-ratio statistics. This trend was similar for both Alexandria and Bemidji as well that had significantly smaller alpha diversity values relative to other WWTPs, whereas Lanesboro was higher than most others. Repeated measure correlations (**Supplementary Figure 2**) of Shannon diversity was significantly, positively correlated with the sum of environmental bacteria abundances (*q* < 0.001, *r_rm_* = 0.21) while Observed diversity was negatively correlated with the sum of human bacteria abundances (*q* = 0.003, *r_rm_* = -0.15), suggesting lower alpha diversity is largely driven by richness of environmental species. Differences in Shannon diversity from most comparisons highlights that each facility has a unique microbiome comprised of both human and environmental bacteria.

**Figure 2.**
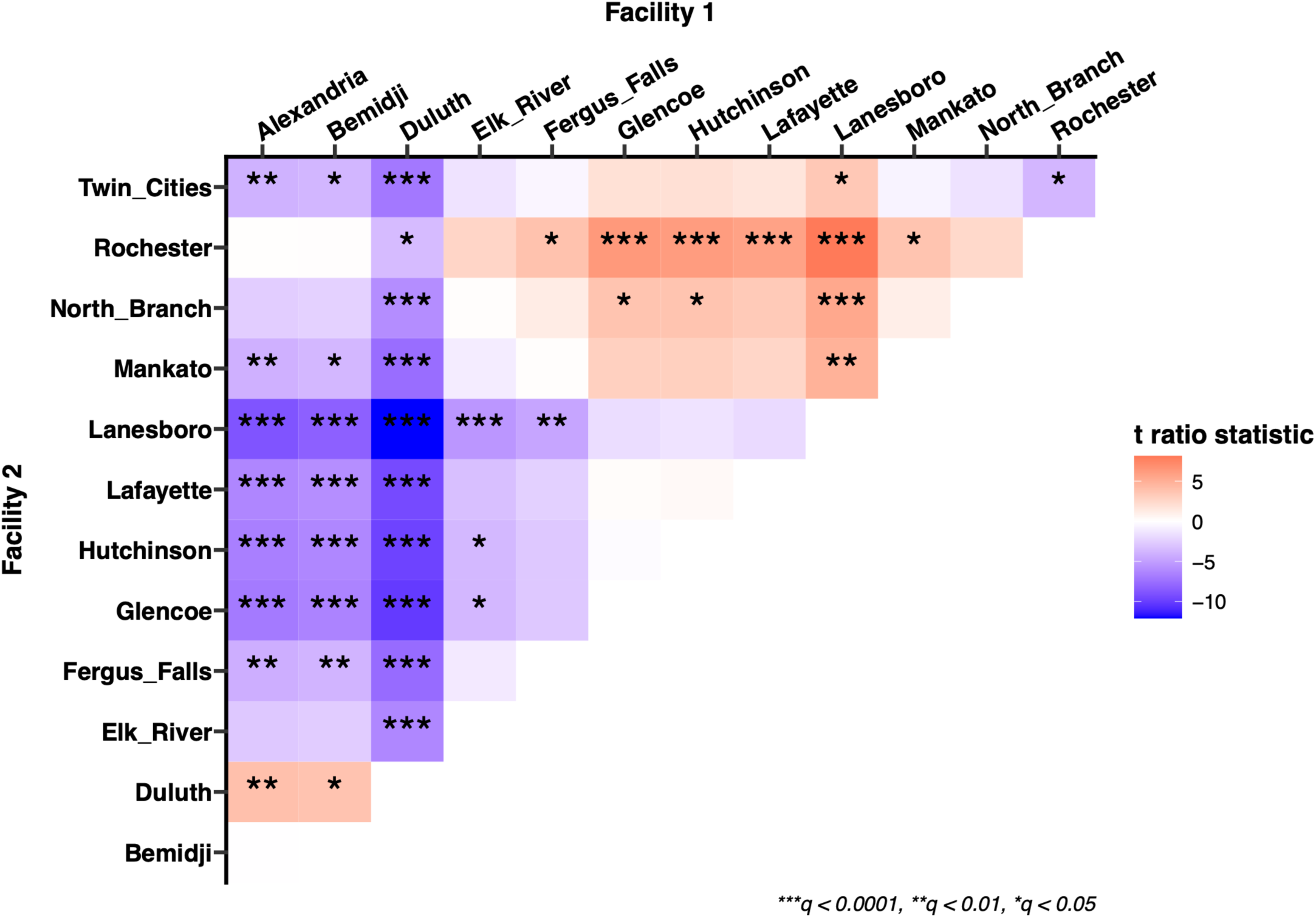
Pairwise comparison of estimated marginal means of alpha diversity between facilities. WWTP alpha diversity was calculated by Shannon Index to quantify the richness and evenness of the microbiome and the predicted mean was compared between facilities and was considered significant with a Tukey-adjusted p-value < 0.05. Positive *t* ratio statistics are red indicating Facility 1 alpha diversity is greater than Facility 2 while negative are blue indicating lesser alpha diversity values from Facility 1 compared to Facility 2. Asterisks denote significance levels of adjusted *p*- values.

A PCA analysis indicated that most facilities are different from one another, however some facilities clustered together (**Figure 3A**). There were significant differences in the clustering of facilities on PCA according to PERMANOVA adjusting for case rate and interacting time (Facility*Collection Week: *p =* 0.029, R-squared = 0.15). A Mantel test with the distance between facility PCA centroids and Haversine distance between Facility locations was not significant (*p* = 0.451, *r* = 0.01) but was more strongly correlated with Bray-Curtis distance centroid Euclidean distances (*p* = 0.18, *r* = 0.16, **Supplementary Figure 3**). An insignificant Mantel test indicates that there is no correlation between these distance measures, suggesting that the PCA clustering observed is not due to geographic proximity or that other factors are more influential such as urbanicity of the surrounding area. The top 10 contributors to PC1 (51.5%) included taxa from the human GI tract and the environment (**Figure 3B**), while PC2 (30.5%) was largely environmental taxa. Altogether, these results suggest that both local environmental and enteric-origin microbes from the residents of the sewer shed region appear to influence wastewater composition to create distinguishable microbiome clusters.

**Figure 3.**
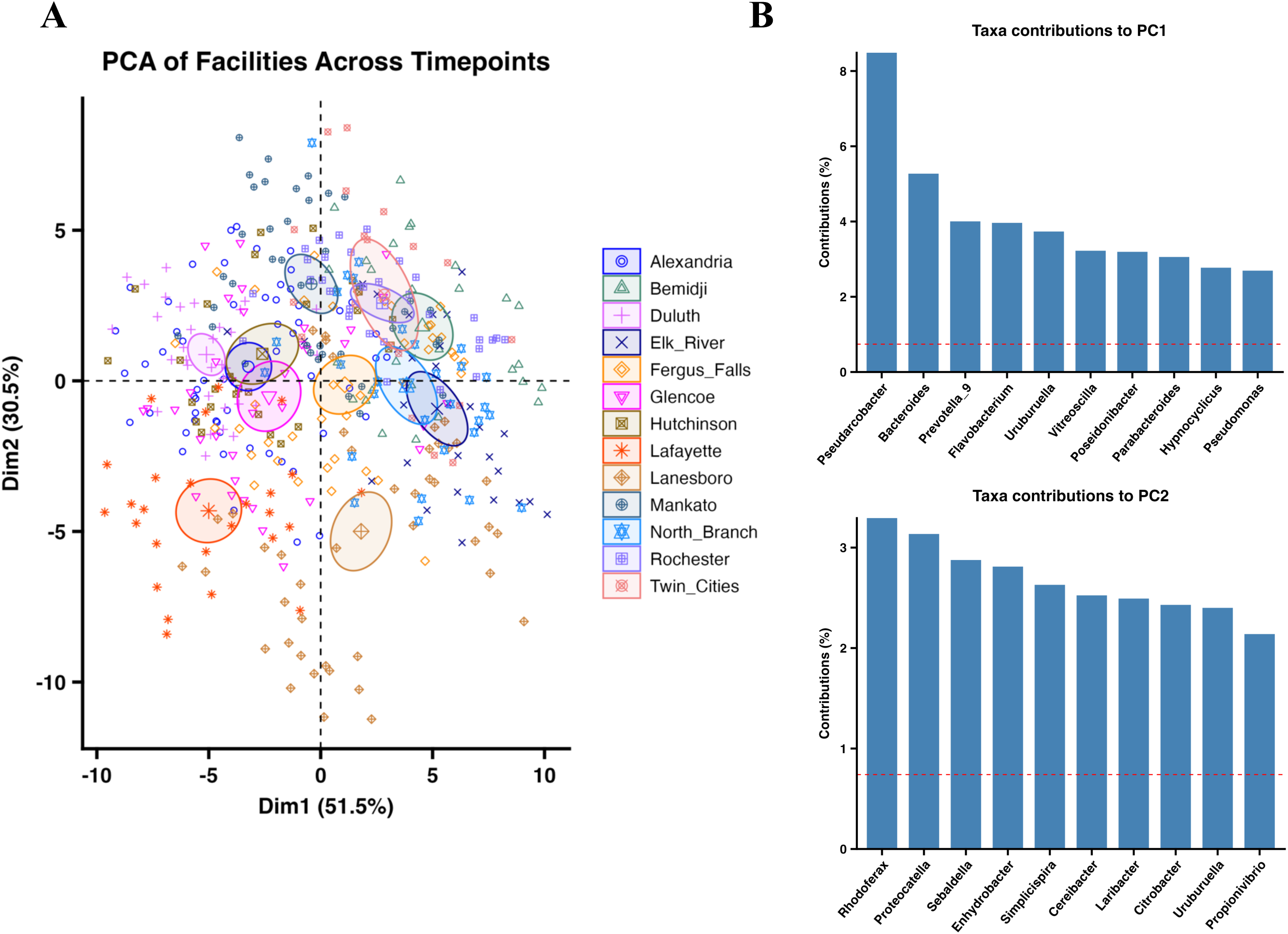
Principal component analysis using individual taxa as contributors. (A) Facilities are represented by separate colors and point shapes with 95% ellipses; centroids for each are matching but larger points. (B) Bar graph scree plots representing the top 10 contributing taxa for PC1 and PC2. The red line indicates the expected average contribution of each variable. Top contributors of PC1 includes enteric bacteria while PC2 is all environmental.

### Wastewater individual taxa are differentially associated with COVID-19 case rate

To determine if specific bacterial taxa in the wastewater were associated with COVID-19 case rate, linear mixed models predicting bacterial relative abundance were constructed while adjusting for weather temperature as a possible confounder. Remarkably, there were 108 taxa that were significantly associated with case rate including both positive and negative relationships after adjusting for temperature (*q* < 0.05, **Figure 4**). Common enteric commensals such as *Blautia, Akkermansia, Subdoligranulum, Lachnospiraceae NK4A136 group, Bifidobacterium, Christensenellaceae R7 group, Anaerostipes, Fusicatenibacter*, and *Dorea* are positively associated with case rate with others being negatively associated including *Bacteroides, Ruminococcus,* and *Parabacteroides.* These genera are core members of the gut microbiome, representing metabolically important producers of metabolites such as short-chain fatty acids and are commonly associated with positive health outcomes^36^.

**Figure 4.**
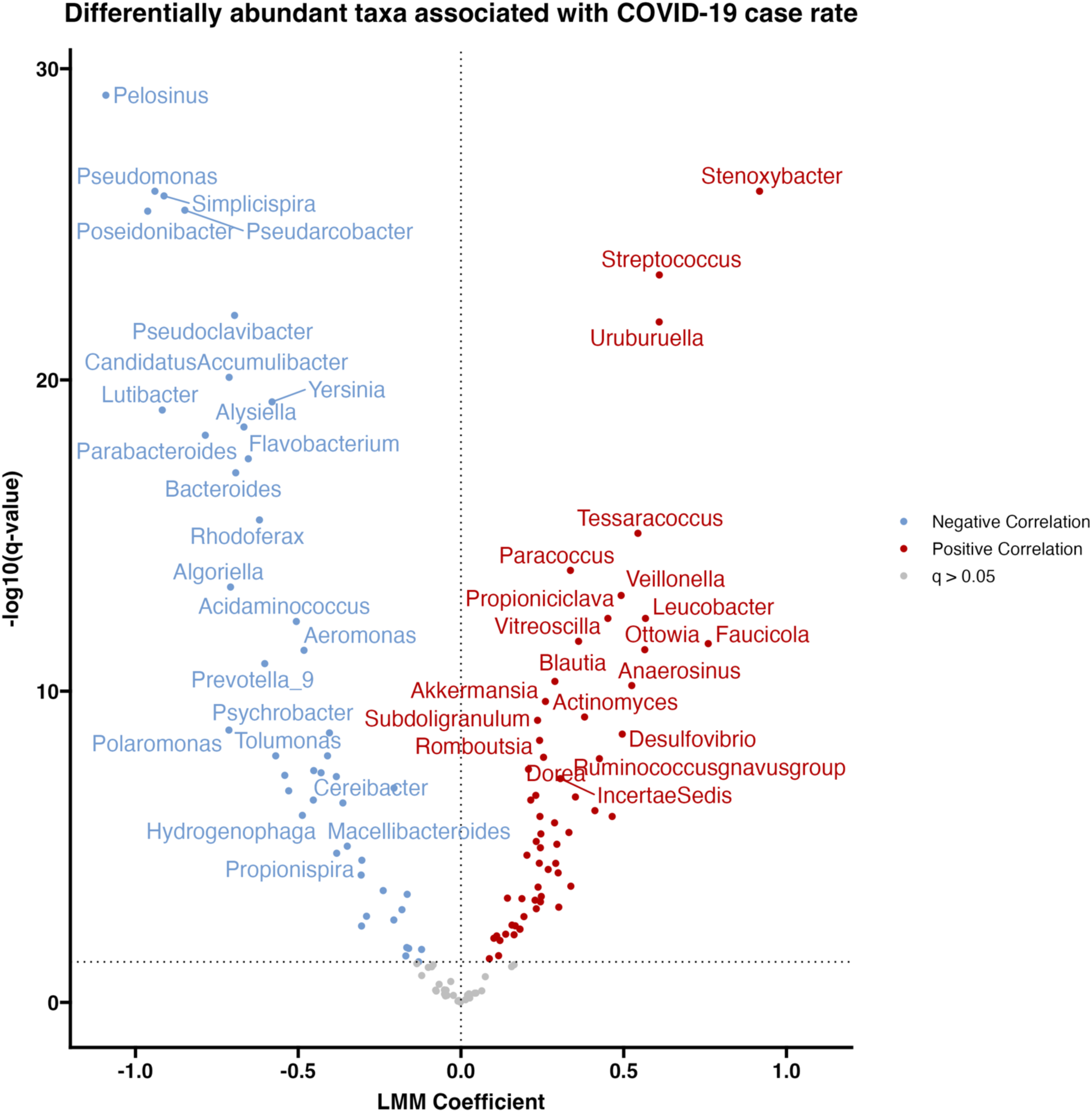
Volcano plot of bacteria associated with case rate. Differential abundance of bacteria associated with COVID-19 case rates after adjusting for temperature where red indicates a positive linear mixed model (LMM) coefficient and blue indicates a negative coefficient. Points in gray did not reach the significance threshold of *q* < 0.05.

Taxa that are more commonly considered to be pathobionts such as *Prevotella* (9) decreased while *Veillonella, Ruminococcus gnavus group*, *Ruminococcus torques group*, *Ruminococcus gavreauii group*, *[Ruminococcaceae] CAG-352, Enterococcus*, *Alistipes*, *Collinsella, Dialister, Streptococcus,* and *Escherichia/Shigella* were positively associated with case rate. Overall, pathogenic taxa were more represented in those associated with case rate compared to commensals, potentially indicating that the disease state that describes COVID-19 infection is perpetuated by an overgrowth of these disease-associated genera. An increased relative abundance of known beneficial bacteria could occur due to an expulsion of bacteria from inflammatory interruption of the gut lining, especially those that reside in the mucosa such as *Akkermansia*.

Several bacteria strongly associated with case rate likely originated from the wastewater itself or from the local environment. Of the top taxa associated with case rate (*Pelosinus, Pseudomonas, Stenoxybacter, Simplicispira, Pseudoarcobacter, Poseidonibacter, Pseudoclavibacter, Uruburuella*, and *Candidatus Accumulibacter, q* < 0.001), most are seldom found in the human microbiome or, in the case of taxa like *Pseudomonas*, are also abundantly found in the environment.

### Microbiota in wastewater are predictive of COVID-19 case rate

We next evaluated the ability of the wastewater microbiome to predict COVID-19 case rates by iteratively performing feature selection on a training set to identify parsimonious sets of bacterial taxa, including potential single taxon predictors, then applying each selected feature set to unseen longitudinal data from a held-out facility across multiple model types for comparison (**Figure 5**). For the model predicting case rate without a lead, selected taxa represent a blend of bacteria that are associated with both enteric and environmental origin (**Figure 5A**). Several of the selected bacteria are commonly associated with the stool microbiome during active COVID- 19 infection where *Ruminococcus gnavus group* and *Ruminococcus torques group* are typically considered pathogenic species related to gastrointestinal upset and *Blautia* as an abundant commensal that is depleted during infection, particularly in severe cases^37^. Non-enteric bacteria are predominately standard wastewater and environmental aquatic dwellers such as *Stenoxybacter* and *Tessaracoccus*. Combining feature-selected taxa with N1 gene copies/mL improved predictive performance slightly relative to either predictor alone (RMSE: 0.44, 0.45, 0.43 and MAE: 0.37, 0.37, 0.34 for N1, feature selected, and combined models, respectively) although ANOVA indicated no significance between model types (*p* = 0.59 and *p* = 0.61 for RMSE and MAE) (**Figure 5B**).

**Figure 5.**
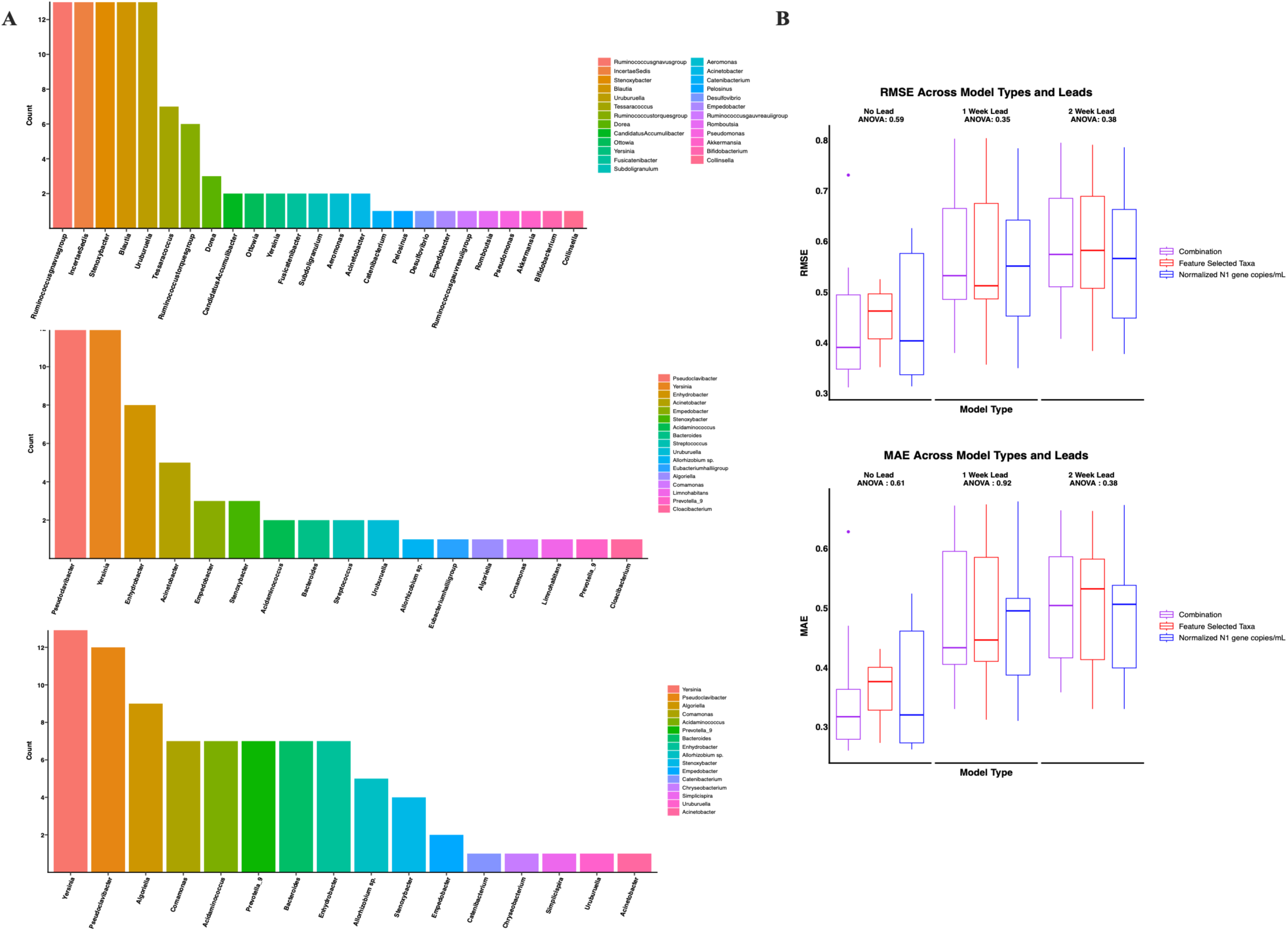
Comparison of models predicting weekly case rates. Model metrics and covariates are shown for no lead (top line), one week lead (middle line), and a two week lead in case rate (bottom line). (A) Barplots showing how frequently specific bacteria were selected during feature selection from glmmLASSO. (B) Boxplots summarizing the RMSE and MAE of predicting case rate on the data of an unseen facility, faceted by

Interestingly, models predicting case rate one week ahead yielded a different selection of bacteria, primarily including environmental bacteria such as *Pseudoclavibacter* and *Yersinia* as the most often selected taxa with only a few enteric genera present such as *Bacteroides*, *Streptococcus*, *Eubacterium halli group,* and *Prevotella 9* (**Figure 5A**). Models including N1 gene copies/mL, feature selected taxa, or a combination of both performed similarly by RMSE and MAE (*p* = 0.35 and *p* = 0.92 for RMSE and MAE, respectively) (**Figure 5B**).

Selected taxa for predicting case rate two weeks ahead was similar to predicting one week ahead (**Figure 5A**). As with models predicting one week ahead, the top two most selected taxa were *Yersinia* and *Pseudoclavibacter*. In terms of model performance, each model again had similar RMSE and MAE (*p* = 0.38 and *p* = 0.38 for RMSE and MAE) (**Figure 5B**). Models tended to work consistently when predicting case rate based on facility, where facilities such as Duluth were predicted worse compared to most others at no lead (**Supplementary Figure 4**).

Overall, incorporating specific members of the microbiome as covariates to predict case rate performed similarly to N1 alone and when used in combination, had modest improvements comparatively highlighting the microbiome’s ability to predict community case rate.

## Discussion

While COVID-19 case rates have decreased from early in the pandemic, COVID-19 remains a continuous burden on the healthcare system and methods such as ours offer increased analytical and correlative resolution. We collected longitudinal wastewater samples from multiple sites across the state of Minnesota to characterize the wastewater microbiome during the COVID-19 pandemic and assess its predictive ability on case rates. When comparing facilities, we observed many differences in alpha diversity metrics as well as distinct microbial compositions that appear to be associated with the size of the sewer shed population and potentially to a lesser degree, geographical proximity, as sites closer to each other tended to be more similar and consistent with previous reports^38,39^. These differences also likely reflect changes in environmental bacteria and abiotic factors such as the surrounding landscape, urban design influence on catchment regions, materials, or treatment protocols of the wastewater facilities.

Differential abundance analysis identified several genera that were strongly associated with COVID-19 case rate. Here, we observed that key gut commensal bacteria such as *Blautia*, *Akkermansia, Anaerostipes, Dorea,* and *Bifidobacterium* were positively associated with case rate. These bacteria are widely recognized to be keystone members that bolster gut health through competition with pathogenic bacteria and modulating the GI metabolic landscape by producing or mediating the production of critical metabolites^40–45^. *Ruminococcus gnavus group*, previously reported to be enriched in severe COVID-19 and associated with GI symptoms and diarrhea^13,46^, was similarly positively associated with case rate. These associations are particularly compelling as they suggest that community-level shifts in the gut microbiome during periods of elevated SARS-CoV-2 transmission, including the shedding of commensal bacteria into wastewater, may serve as a detectable and informative signal of COVID-19 burden. While other respiratory illnesses are known to impact stool bacterial compositions, social behavior during the study collection period (i.e. contact-tracing, self-isolating protocols, and social distancing) resulted in a unique absence of other respiratory viruses limiting the impact of these other illnesses on the wastewater microbiome^47^. Further, these findings were strongly significant despite adjusting for temperature, suggesting that seasonality is not the reason for the observed relationship but instead that the non-enteric bacteria are influenced by disease pathogenesis from acute COVID-19 infection.

We expanded upon these associations between microbial taxa in wastewater and COVID- 19 case rates using predictive modeling, feature selection, and cross validation to evaluate the ability of the wastewater microbiome at estimating COVID-19 county-level case rate and compare to SARS-CoV-2 N1 gene copies/mL^19^. Models were evaluated using RMSE and MAE for their predictive error and tested using COVID-19 case rates up to a two-week lead time.

When estimating the present week’s case rate from covariates, a combined model using both N1 genes copies and feature selected taxa performed slightly better. Feature selected taxa and N1 models likely performed similarly, especially at predicting future case rates, since SARS-CoV-2 viral load peak is typically aligned with symptom onset^48^ and variably persists up to months after respiratory clearance^49^ but may indicate the infection capacity of the population, whereas the microbiome may be influenced earlier than respiratory symptom occurrence^50^. Feature selection appeared to identify bacteria that may be associated with different stages of infection depending on which week was being predicted. For example, using the same week’s case rate as the dependent variable resulted in higher representation of bacteria commonly associated with COVID-19 GI symptoms (*Ruminococcus gnavus group, Blautia, Dorea,* and *Ruminococcus torques group*^51–54^. This week had the greatest diversity in bacteria selected with most being enteric, indicating the magnitude of impact COVID-19 infection has on the gut microbiome.

For predicting case rates 1 or 2 weeks ahead, feature selected taxa became more represented by environmental taxa, but bacteria still frequently associated with COVID-19, specifically *Prevotella 9* (mainly comprised of *P. copri*), *Bacteroides,* and *Streptococcus* which has been associated with extended viral shedding in COVID-19 infection^55^. Numerous environmental taxa were included by features selection such as *Yersinia, Pseudoclavibacter,* and *Stenoxybacter* and, when used in models, performed similarly to models comprised of direct measures of viral shedding (N1). The relationship between these taxa and COVID-19 cases is, to our knowledge, previously unreported, however, other environmental taxa such as *Aeromonas* have been observed^17^ which was identified to a lesser extent in feature selection. Alterations of the input into wastewater, both in the taxa and the stool metabolome^56,57^, changes the competitive landscape on a previously stable system^58^ where certain bacteria may be enriched or inhibited. In addition, due to the compositional nature of the data, environmental bacteria may act as a proxy for more subtle enrichment in several rarer human taxa in wastewater with small effect sizes that would not have been selected for prediction or potentially been removed by filtering thresholds. Altogether, the wastewater microbiome offers a rich pool of data that is useful in prediction of COVID-19 cases alongside previously used surveillance markers. Future work should test similar methods for other infectious diseases that impact the gut microbiome and pose a large public health burden^59–61^, especially in regions with underfunded health care services that would disproportionately benefit from improved wastewater surveillance.

Altogether, the wastewater microbiome offers a rich pool of data that is useful in prediction of COVID-19 cases and is additive to other employed surveillance markers. The serial analysis of a diverse, pooled population with fluctuating infection rates and evolving microbial consistency warrants the detailing of potential limitations. First, the finest resolution available for community case rate was at the county level, despite facilities typically serving a smaller catchment area. Despite less travel overall during this period, we could not account for movement between populations such as commuters and asymptomatic or mild cases which likely contribute to both SARS-CoV-2 gene copies and altered microbiome input into wastewater.

Further, microbiome analysis was restricted to compositional genus level data, further information could be gleaned by species or strain level taxonomic resolution from increased sequencing depth or by other sequencing strategies. Finally, our predictive models were limited to prediction respective to an individual facility and training on a set of twelve for cross validation. Despite these limitations, this analysis still effectively demonstrates the utility of microbiome data in wastewater-based surveillance.

Our study offers significant insights into wastewater bacterial relationships with COVID- 19 community case rates and its usefulness in predicting future infections. We observed many taxa that exhibit strong relationships with COVID-19, many of which are reportedly altered during infection. We applied the microbiome data to predictive modeling and demonstrated that the microbiome could effectively estimate case rate in present and future weeks on par with previously employed public health methods. Further, the taxa selected with different leads using this technique identified trends potentially related to separate stages of infection. Following similar modeling approaches may prove useful in wastewater surveillance for ongoing SARS-CoV-2 variants, long COVID prevalence tracking, and holds tremendous potential for offering a method for tracking and associating other infectious diseases with public health mitigation efforts.

## Data Availability

Data available on Sequence Read Archive.

## ABBREVIATIONS

16S rRNA V4: 16S ribosomal ribonucleic acid variable region 4
ACE2: Angiotensin converting enzyme 2
ANOVA: Analysis of variance
ASVs: Amplicon sequence variants
BIC: Bayesian information criterion
CLR: Center log ratio
COVID-19: Coronavirus disease 2019
DNA: Deoxyribonucleic acid
Emmeans: Estimated marginal means
GI: Gastrointestinal
glmmLASSO: Generalized linear mixed models least absolute shrinkage operator
LGOCV: Leave-group-out cross validation
MAE: Mean absolute error
MDH: Minnesota Department of Health
N1/N2: Nucleocapsid 1 or 2 (gene of SARS-CoV-2)
NOAA: National Oceanic Atmospheric Administration
NP: Nasopharyngeal
OP: Oropharyngeal
PCA: Principal component analysis
PCoA: Principal coordinate analysis
PERMANOVA: Permutation analysis of variance
qRT-PCR: Quantitative reverse-transcriptase polymerase chain reaction
RCLR: Robust center log ratio
RMSE: Root mean squared error
RT-PCR: Reverse-transcriptase polymerase chain reaction
SARS-CoV-2: Severe acute respiratory syndrome coronavirus 2
SCFA: Short-chain fatty acid

## DECLARATIONS

### Consent for publication

Not applicable.

### Availability of data and materials

Raw data sequences are available on Sequence Read Archive (SRA) with the accession number PRJNA1511278. Code is available upon request.

### Author contributions

Conceptualization: NRK, TAS, CMB, MJO, TWS

Supervision: NRK

Funding acquisition: MJO, TWS, NRK

Investigation: TAS, CMB, MB, ES, CB

Formal analysis: TAS

Visualization: TAS

Writing – original draft: TAS

Writing – review & editing: NRK, CMB, ES, MB, SRC, AV, JL, CB, TWS, MJO

## Acknowledgements

Microbiome 16S sequencing was conducted by the University of Minnesota Genomics Center.

