## Supplemental Figure 1 for "Microbiome signature in wastewater is predictive of COVID-19 case rates"

**SUPPLEMENTARY**

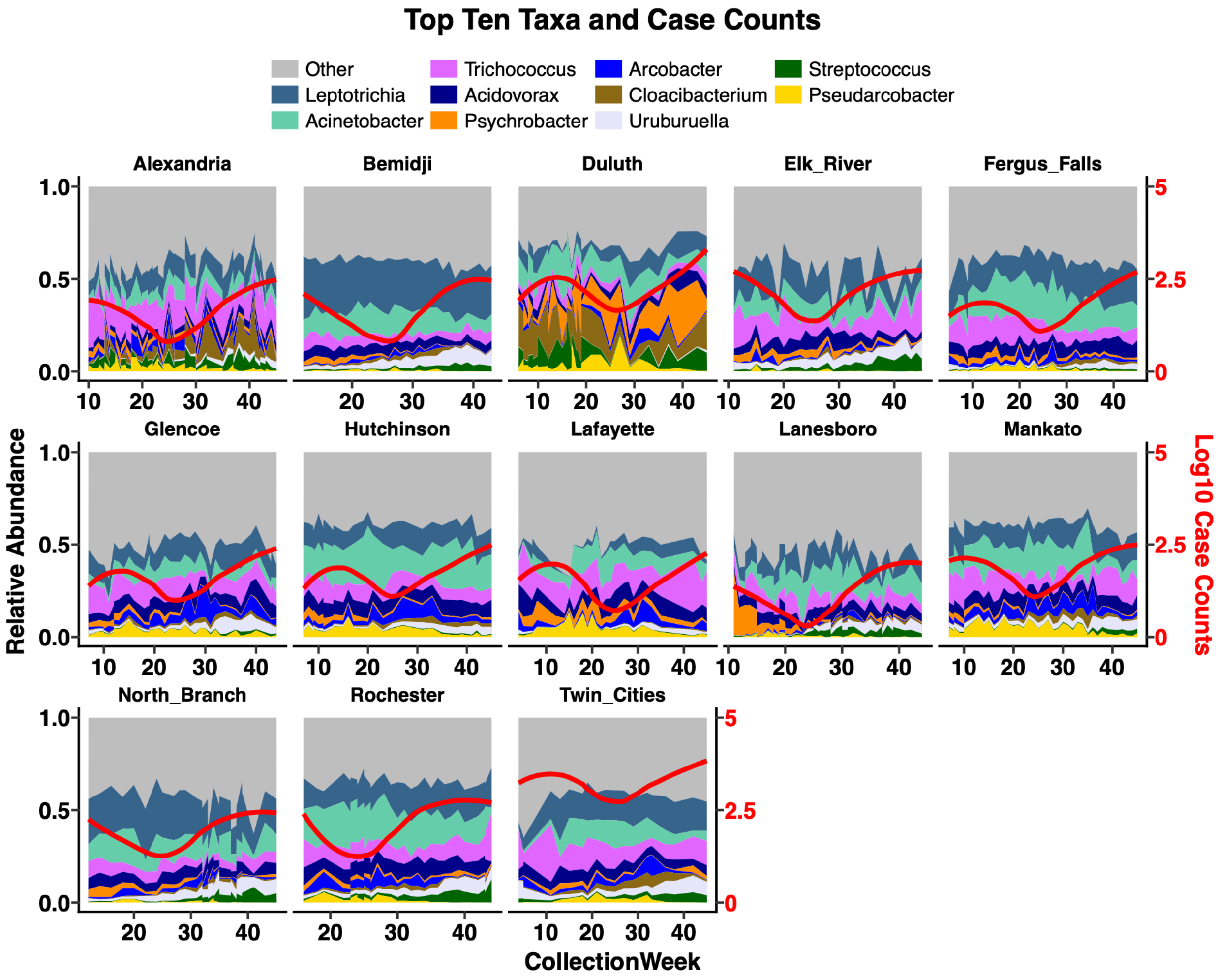


**Supplementary Figure 1. Longitudinal area plots of each facility.** Area plots feature relative abundance of all bacteria over time, with the top 10 taxa assigned individual colors. All other taxa are included in the Other category. Red lines represented log10 county case count loess fits for each facility.
