## Supplemental Figure 2 for "Microbiome signature in wastewater is predictive of COVID-19 case rates"

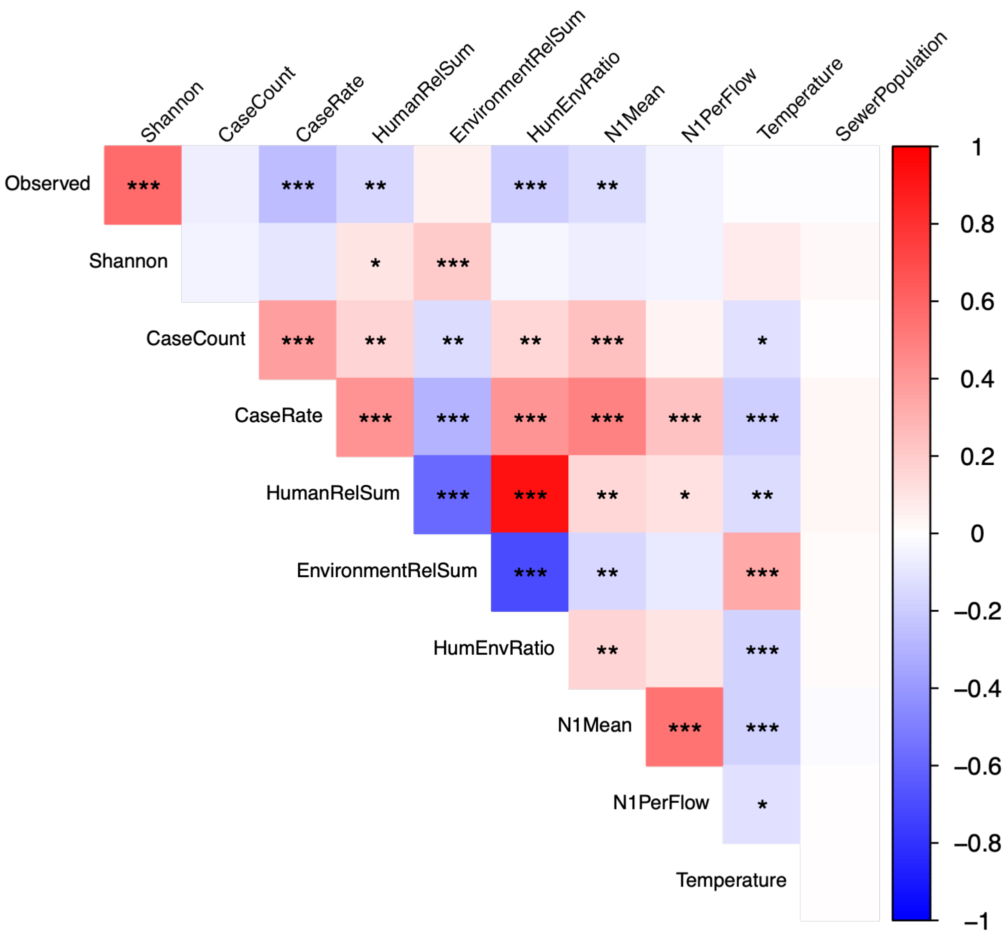


**Supplementary Figure 2. Repeated measure correlations between study covariates.** Heatmap representing correlations between study covariates where warm colors represent positive association and blue indicate negative. Asterisks denote significance levels of 0.05, 0.01, or 0.001for *, **, or *** respectively.
