## Supplemental Figure 3 for "Microbiome signature in wastewater is predictive of COVID-19 case rates"

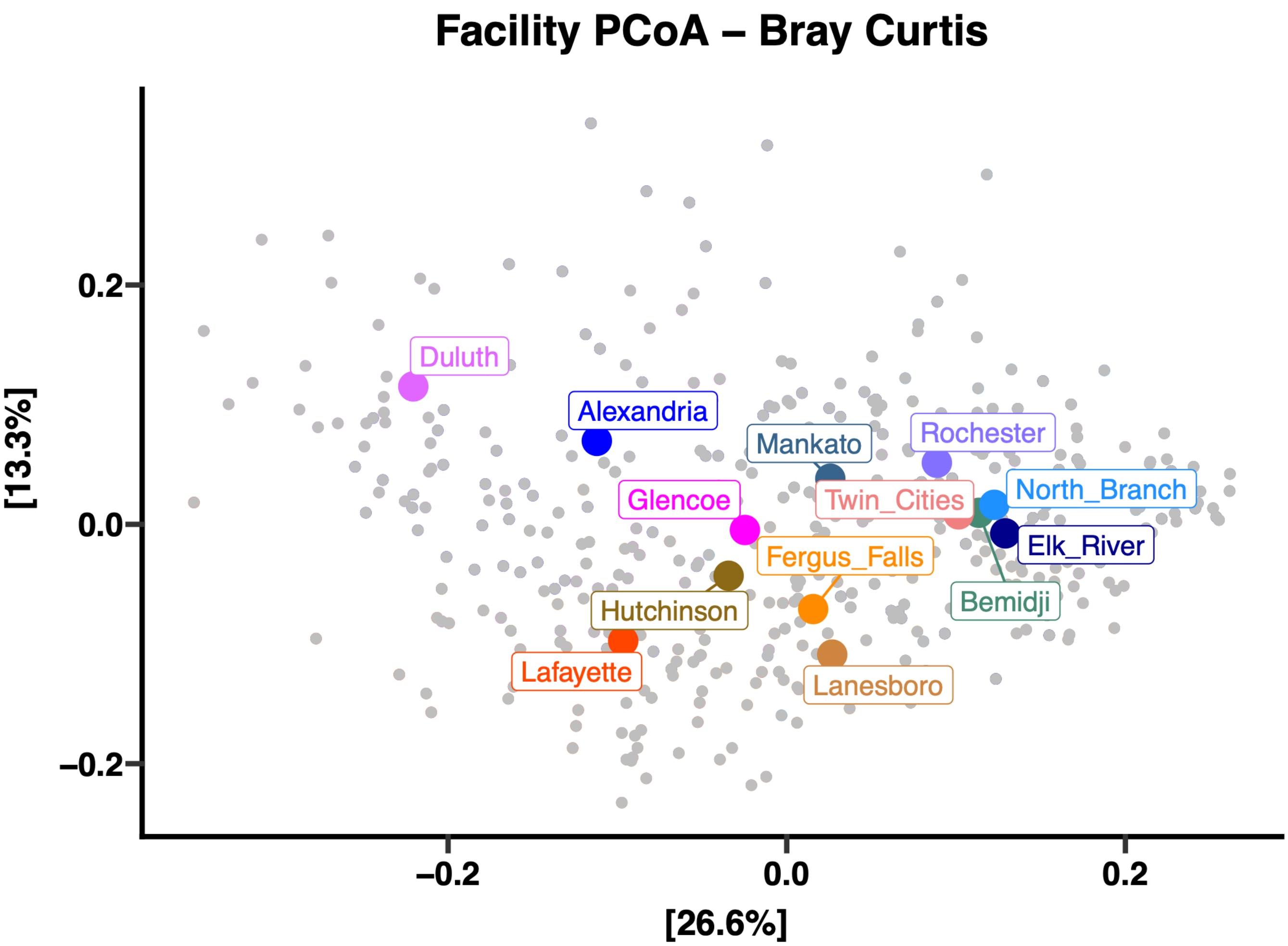


**Supplementary Figure 3. Principal coordinate analysis using Bray-Curtis dissimilarity between WWTPs.** Bray-Curtis distance was calculated between each sample to describe their relative dissimilarity in microbiome composition. Percent on the axes describes the amount of variance each explains. Colored points are centroids of all collected samples from a facility which are labeled with their own colors.
