## Supplemental Figure 4 for "Microbiome signature in wastewater is predictive of COVID-19 case rates"

**A**

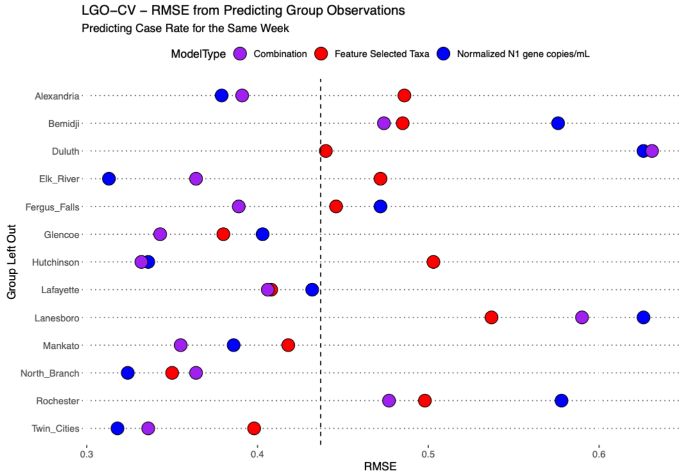


**B**

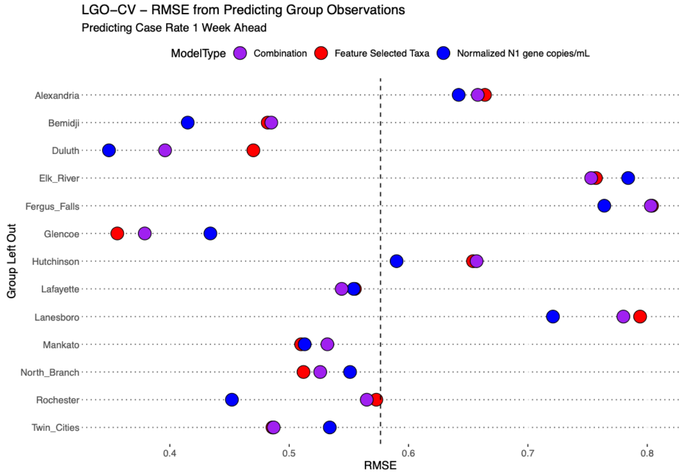


**C**

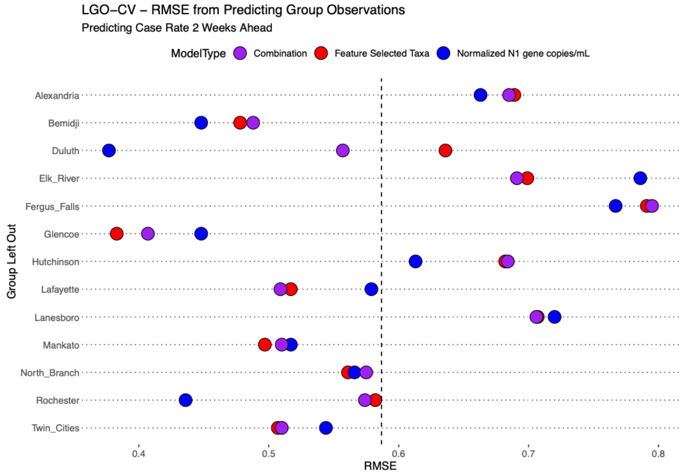


**Supplementary Figure 4. Dot plots of model performance on test data.** Plots show model performance by RMSE for (A) no lead, (B) 1 week lead, and (C) 2 week lead. Vertical dotted lines indicate the average RMSE across all models. WWTPs on y-axis represent the test group for each model. Color of dots represent the model type with red for feature selected taxa, blue for N1, and purple for a combination.
